# Unreported cases for Age Dependent COVID-19 Outbreak in Japan

**DOI:** 10.1101/2020.05.07.20093807

**Authors:** Quentin Griette, Pierre Magal, Ousmane Seydi

## Abstract

We investigate the age structured data for the COVID-19 outbreak in Japan. We consider a mathematical model for the epidemic with unreported infectious patient with and without age structure. In particular, we build a new mathematical model and a new computational method to fit the data by using age classes dependent exponential growth at the early stage of the epidemic. This allows to take into account differences in the response of patients to the disease according to their age. This model also allows for a heterogeneous response of the population to the social distancing measures taken by the local government. We fit this model to the observed data and obtain a snapshot of the effective transmissions occurring inside the population at different times, which indicates where and among whom the disease propagates after the start of public mitigation measures.

## 1 Introduction

COVID-19 disease caused by the corona virus SARS-CoV-2 first appeared in Wuhan, China, and the first cases were notified to WHO on December 31, 2019 [33, 34]. Beginning in Wuhan as an epidemic, it then spread very quickly around the world to become a global pandemic on March 11, 2020 [33]. Symptoms of this disease include fever, shortness of breath, cough, and a non-negligible proportion of infected individuals may develop severe forms of the symptoms leading to their transfer to intensive care units and, in some cases, death, see *e.g*. Guan et al [8] and Wei et al [27]. Both symptomatic and asymptomatic individuals can be infectious [22, 27, 30], which makes the control of the disease particularly challenging.

The virus is characterized by its rapid progression among individuals, most often exponential in the first phase, but also a marked heterogeneity in populations and geographic areas [31, 29, 26]. The number of reported cases worldwide exceeded 3 millions as of May 3, 2020 [32]. The heterogeneity of the number of cases and the severity according to the age groups, especially for children and elderly people, aroused the interest of several researchers [4, 20, 16, 23, 24]. Indeed, several studies have shown that the severity of the disease increases with the age and co-morbidity of hospitalized patients (see *e.g*. To et al [24] and Zhou et al [29]). Wu et al [28] have shown that the risk of developing symptoms increases by 4% per year in adults aged between 30 and 60 years old while Davies et al [5] found that there is a strong correlation between chronological age and the likelihood of developing symptoms. Since completely asymptomatic individuals can also be contagious, a higher probability of developing symptoms does not necessarily imply greater infectiousness: Zou et al [30] found that, in some cases, the viral load in asymptomatic patients was similar to that in symptomatic patients. Moreover while adults are more likely to develop symptoms, Jones et al [9] found that the viral loads in infected children do not differ significantly from those of adults.

These findings suggest that a study of the dynamics of inter-generational spread is fundamental to better understand the spread of the corona virus and most importantly to efficiently fight the COVID-19 pandemic. To this end the distribution of contacts between age groups in society (work, school, home, and other locations) is an important factor to take into account when modeling the spread of the epidemic. To account for these facts, some mathematical models have been developed [1, 3, 5, 20, 23]. In Ayoub et al [1] the authors studied the dependence of the COVID-19 epidemic on the demographic structures in several countries but did not focus on the contacts distribution of the populations. In [3, 5, 20, 23] a focus on the social contact patterns with respect to the chronological age has been made by using the contact matrices provided in Prem et al [19]. While Ayoub et al [1], Chikina and Pegden [3] and Davies et al [5] included the example of Japan in their study, their approach is significantly different from ours. Indeed, Ayoub et al [1] use a complex mathematical model to discuss the influence of the age structure on the infection in a variety of countries, mostly through the basic reproduction number 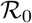. They use parameter values from the literature and from another study of the same group of authors [2], where the parameter identification is done by a nonlinear least-square minimization. Chikina and Pegden [3] use an age-structured model to investigate age-targeted mitigation strategies. They rely on parameter values from the literature and do discuss using age-structured temporal series to fit their model. Finally, Davies et al [5] also discuss age-related effects in the control of the COVID epidemic, and use statistical inference to fit an age-structured SIR variant to data; the model is then used to discuss the efficiency of different control strategies. We provide a new, explicit computational solution for the parameter identification of an age-structured model. The model is based on the SIUR model developed in Liu et al [11], which accounts for a differentiated infectiousness for reported and non-reported cases (contrary to, for instance, other SIR-type models). In particular, our method is significantly different from nonlinear least-squares minimization and does not involve statistical inference.

In this article we focus on an epidemic model with unreported infectious symptomatic patients (i.e. with mild or no symptoms). Our goal is to investigate the age structured data of the COVID-19 outbreak in Japan. In section 2 we present the age structured data and in section 3 the mathematical models (with and without age structure). One of the difficulties in fitting the model to the data is that the growth rate of the epidemic is different in each age class, which lead us to adapt our early method presented in Liu et al [11]. The new method is presented in the Appendix A. In section 4 we present the comparison of the model with the data. In the last section we discuss our results.

## 2 Data

Patient data in Japan have been made public since the early stages of the epidemic with the quarantine of the *Diamond Princess* in the Haven of Yokohama. We used data from the website covid19japan.com^1^ which is based on reports from national and regional authorities. Patients are labeled “confirmed” when tested positive to COVID-19 by PCR. Interestingly, the age class of the patient is provided for 13 660 out of 13970 confirmed patients (97.8% of the confirmed population) as of April 29. The age distribution of the infected population is represented in Figures 1 compared to the total population per age class (data from the Statistics Bureau of Japan estimate for October 1, 2019). In Figure 2 we plot the number of reported cases per 10 000 people of the same age class (*i.e*. the number of infected patients divided by the population of the age class times 10 000). Both datasets are given in Table 1 and a statistical summary is provided by Table 2. Note that the high proportion of 20-60 years old confirmed patients may indicate that the severity of the disease is lower for those age classes than for older patients, and therefore the disease transmits more easily in those age classes because of a higher number of asymptomatic individuals. Elderly infected individuals might transmit less because they are identified more easily. The cumulative number of death (Figure 5) is another argument in favor of this explanation. We also reconstructed the time evolution of the reported cases in Figure 3 and Figure 4. Note that the steepest curves precisely concern the 20-60-years old, probably because they are economically active and therefore have a high contact rate with the population.

**Figure 1:**
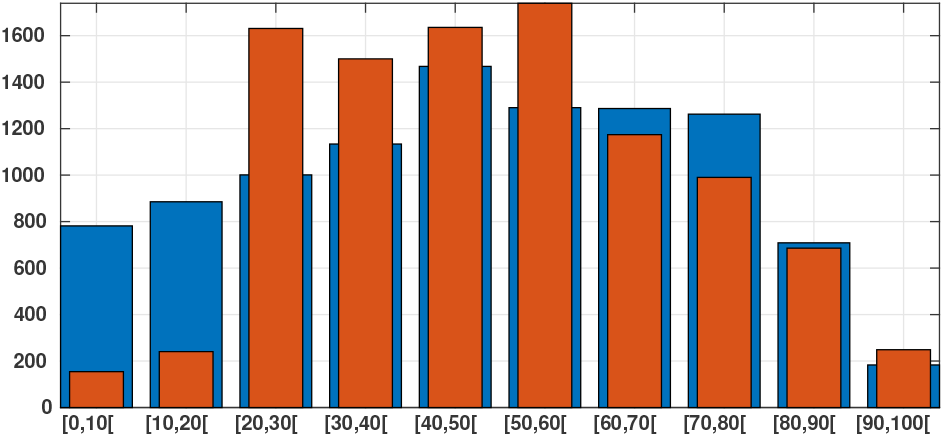
*In this figure we plot in blue bars the age distribution of the Japanese population for* 10 000 *people and we plot in orange bars the age distribution of the number of reported cases of SARS-CoV-2 for* 10 000 *patient on April 29 (based on the total of* 13660 *reported cases). We observe that* 77% *of the confirmed patients belong to the 20–60 years age class*.

**Figure 2:**
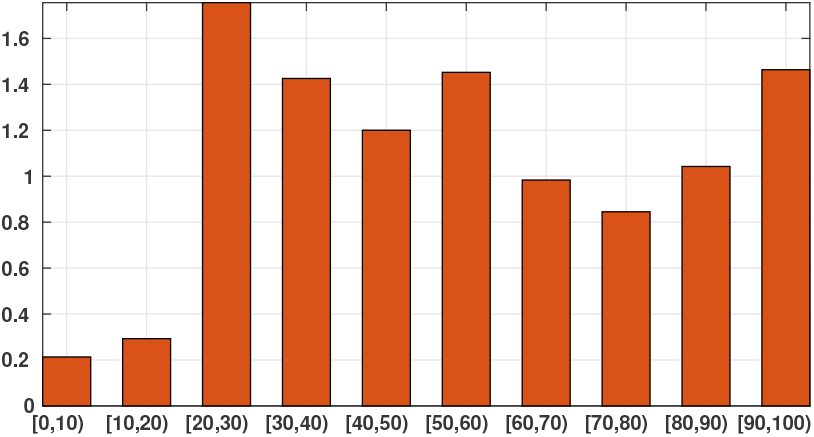
*In this figure we plot the number of infected patients for each age class per* 10 000 *individuals of the same age class (i.e. the number of infected individuals divided by the population of the age class times* 10 000*). The figure shows that the individuals are more or less likely to becomes infected depending on their age class. The bars describe the susceptibility of people to the SARS-CoV-2 depending on their age class*.

**Figure 3:**
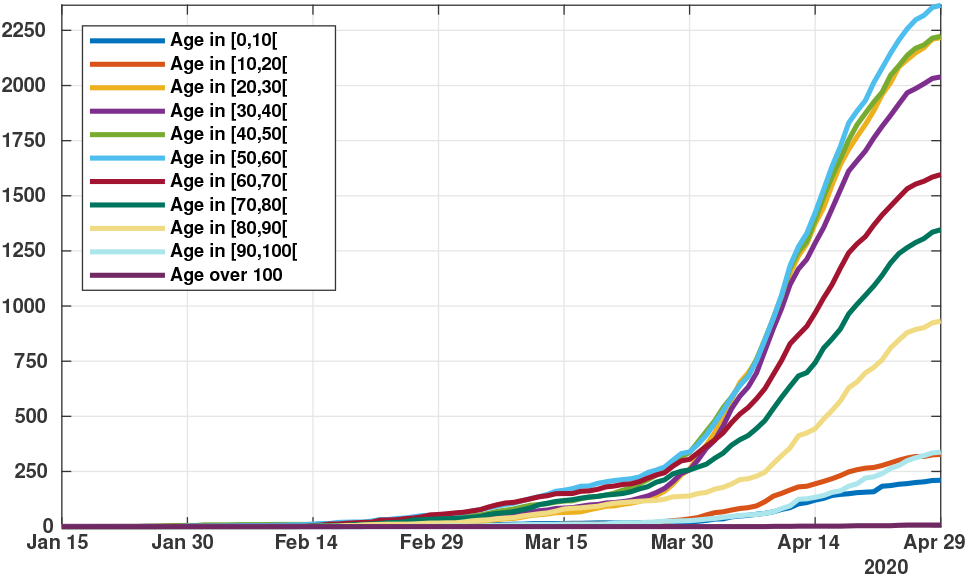
*Time evolution of the cumulative number of reported cases of SARS-CoV-2 per age class. The vertical axis represents the total number of cumulative reported cases in each age class*.

**Figure 4:**
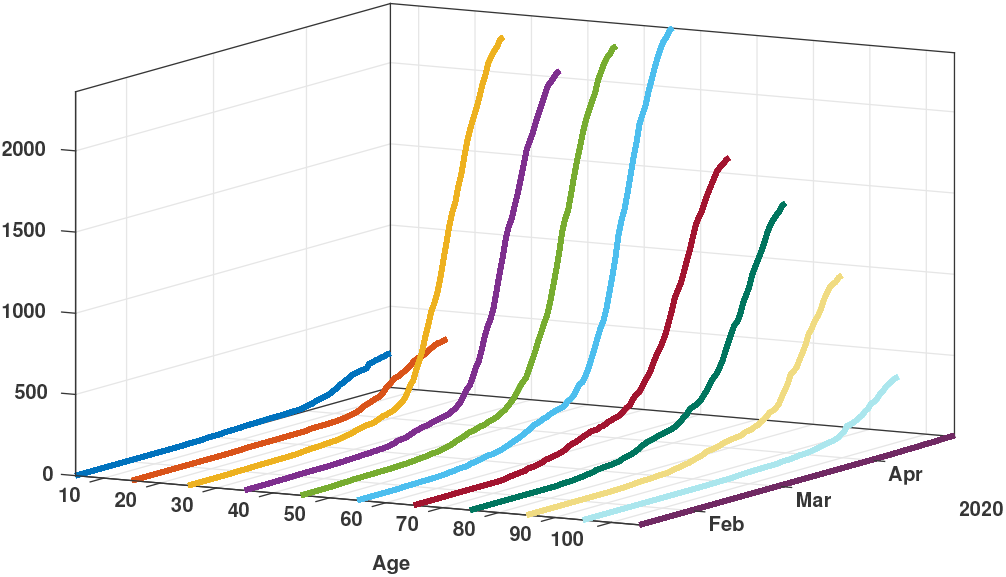
*Time evolution of the cumulative number of reported cases of SARS-CoV-2 per age class. The vertical axis represents the total number of cumulative reported cases in each age class*

**Figure 5:**
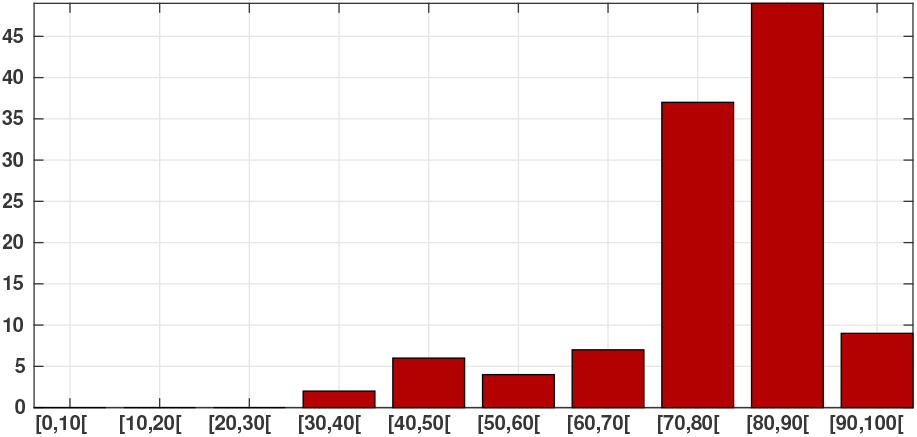
*Cumulative number of SARS-CoV-2-induced deaths per age class (red bars). We observe that* 83% *of death occur in between* 70 *and* 100 *years old*.

**Table 1:**
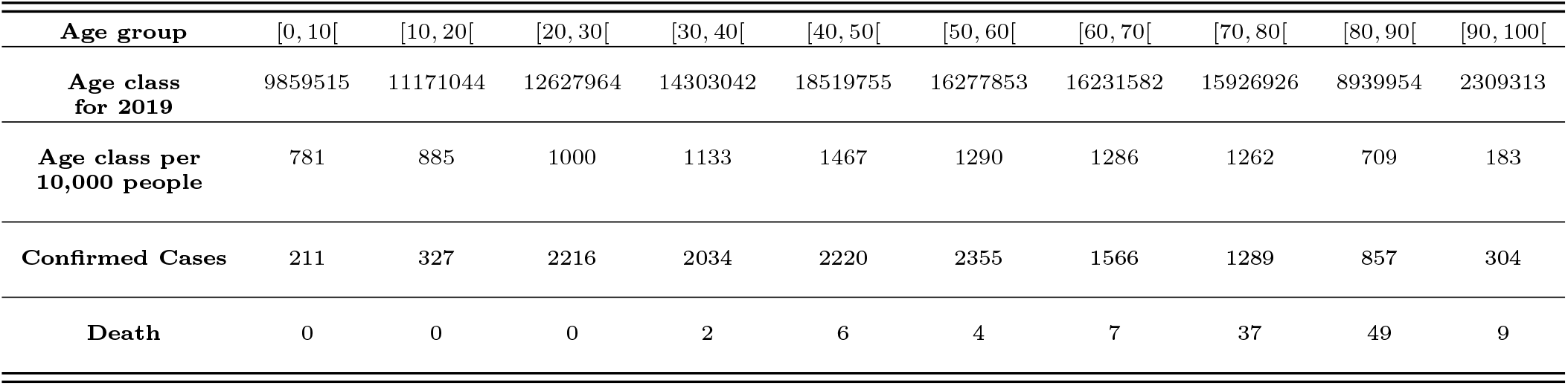
*The age distribution of Japan is taken from the Statistics Bureau of Japan [35]. The number of cases and the number of death the data come from Prefectural Governments and Japan Ministry of Health, Labour and Welfare*.

**Table 2:** *Statistical summary of the data from Table 1*.

| Dataset | Japanese population | Infected | Deceased |
| --- | --- | --- | --- |
| First Quartile | 28 | 28 | 68 |
| Median | 48 | 44 | 75 |
| Third Quartile | 67 | 59 | 81 |

## 3 Methods

### 3.1 SIUR Model

The model consists of the following system of ordinary differential equations:

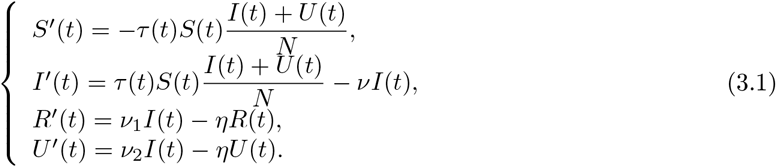

This system is supplemented by initial data

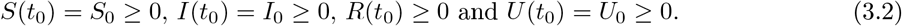

Here *t* ≥ *t*_0_ is time in days, *t*_0_ is the starting date of the epidemic in the model, *S*(*t*) is the number of individuals susceptible to infection at time *t*, *I*(*t*) is the number of asymptomatic infectious individuals at time *t*, *R*(*t*) is the number of reported symptomatic infectious individuals at time *t*, and *U*(*t*) is the number of unreported symptomatic infectious individuals at time t. A flow chart of the model is presented in Figure 6.

Asymptomatic infectious individuals *I*(*t*) are infectious for an average period of 1/*ν* days. Reported symptomatic individuals *R*(*t*) are infectious for an average period of 1/*η* days, as are unreported symptomatic individuals *U*(*t*). We assume that reported symptomatic infectious individuals *R*(*t*) are reported and isolated immediately, and cause no further infections. The asymptomatic individuals *I*(*t*) can also be viewed as having a low-level symptomatic state. All infections are acquired from either *I*(*t*) or *U*(*t*) individuals. A summary of the parameters involved in the model is presented in Table 3.

Our study begins in the second phase of the epidemics, *i.e*. after the pathogen has succeeded in surviving in the population. During this second phase *τ*(*t*) ≡ *τ*_0_ is constant. When strong government measures such as isolation, quarantine, and public closings are implemented, the third phase begins. The actual effects of these measures are complex, and we use a time-dependent decreasing transmission rate τ(*t*) to incorporate these effects. The formula for *τ*(*t*) is

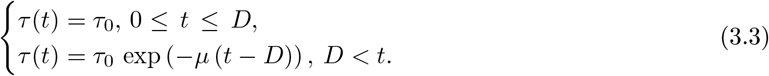

The date *D* is the first day of public intervention and *µ* characterises the intensity of the public intervention.

A similar model has been used to describe the epidemics in mainland China, South Korea, Italy, and other countries, and give reasonable trajectories for the evolution of the epidemic based on actual data [7, 11, 12, 13, 14, 15]. Compared with these models, we added a scaling with respect to the total population size *N*, for consistency with the age-structured model (3.12). This only changes the value of the parameter *τ* and does not impact the qualitative or quantitative behavior of the model.

**Table 3:** *Parameters of the model*.

| Symbol | Interpretation | Method |
| --- | --- | --- |
| $t_0$ | Time at which the epidemic started | fitted |
| $S_0$ | Number of susceptible at time $t_0$ | fixed |
| $I_0$ | Number of asymptomatic infectious at time $t_0$ | fitted |
| $U_0$ | Number of unreported symptomatic infectious at time $t_0$ | fitted |
| $\tau(t)$ | Transmission rate at time $t$ | fitted |
| $D$ | First day of public intervention | fitted |
| $\mu$ | Intensity of the public intervention | fitted |
| $1/\nu$ | Average time during which asymptomatic infectious are asymptomatic | fixed |
| $f$ | Fraction of asymptomatic infectious that become reported symptomatic infectious | fixed |
| $\nu_1 = f\nu$ | Rate at which asymptomatic infectious become reported symptomatic | fixed |
| $\nu_2 = (1 - f)\nu$ | Rate at which asymptomatic infectious become unreported symptomatic | fixed |
| $1/\eta$ | Average time symptomatic infectious have symptoms | fixed |

**Figure 6:**
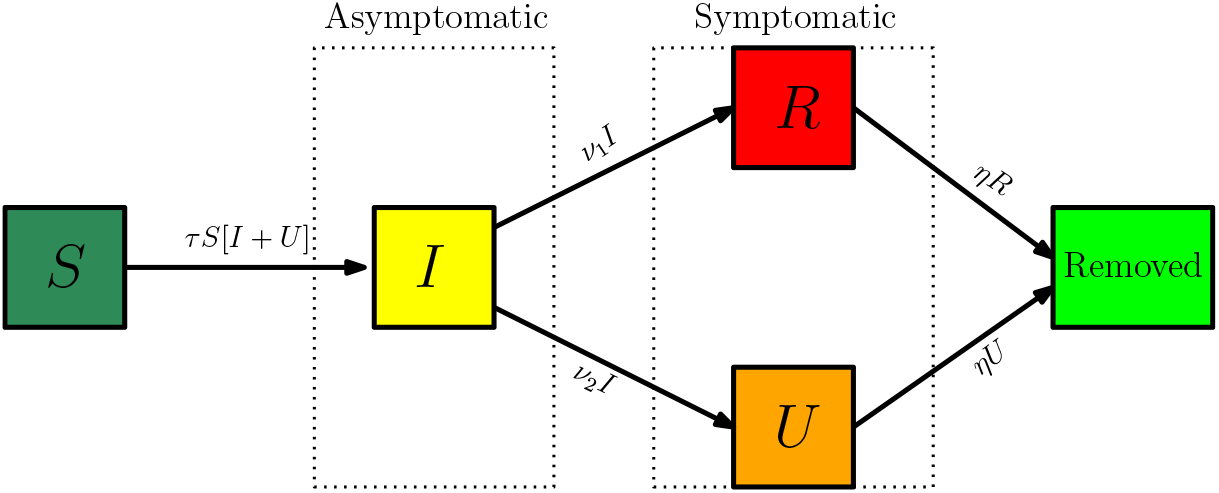
*Compartments and flow chart of the model*.

### 3.2 Comparison of the model (3.1) with the data

At the early stages of the epidemic, the infectious components of the model *I*(*t*), *U*(*t*) and *R*(*t*) must be exponentially growing. Therefore, we can assume that

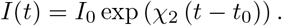

The cumulative number of reported symptomatic infectious cases at time *t*, denoted by *CR*(*t*), is

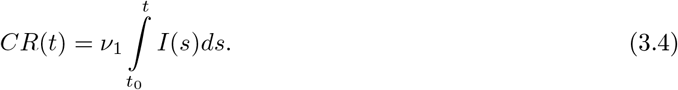

Since *I*(*t*) is an exponential function and *CR*(*t*_0_)=0 it is natural to assume that *CR*(*t*) has the following special form:

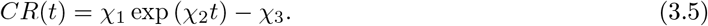

As in our early articles [11, 12, 13, 14, 15], we fix χ_3_ =1 and we evaluate the parameters χ_1_ and χ_2_ by using an exponential fit to

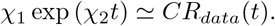

We use only early data for this part, from day *t* = *d*_1_ until day *t* = *d*_2_, because we want to catch the exponential growth of the early epidemic and avoid the influence of saturation arising at later stages.

Remark 3.1

*The estimated parameters* χ_1_ *and* χ_2_ *will vary if we change the interval* [*d*_1_,*d*_2_].

Once χ_1_,χ_2_,χ_3_ are known, we can compute the starting time of the epidemic *t*_0_ from (3.5) as:

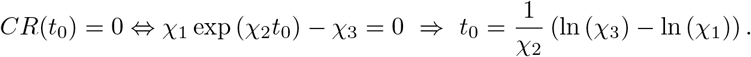

We fix *S*_0_ = 126.8 × 10^6^, which corresponds to the total population of Japan. The quantities *I*_0_, *R*_0_, and *U*_0_ correspond to the values taken by *I*(*t*), *R*(*t*) and *U*(*t*) at *t* = *t*_0_ (and in particular *R*_0_ should not be confused with the basic reproduction number 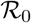). We fix the fraction *f* of symptomatic infectious cases that are reported. We assume that between 80% and 100% of infectious cases are reported. Thus, *f* varies between 0.8 and 1. We assume that the average time during which the patients are asymptomatic infectious 1/*ν* varies between 1 day and 7 days. We assume that the average time during which a patient is symptomatic infectious 1/*η* varies between 1 day and 7 days. In other words we fix the parameters f, *ν, η*. Since f and ν are known, we can compute

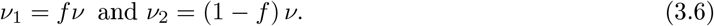

Computing further (see below for more details), we should have

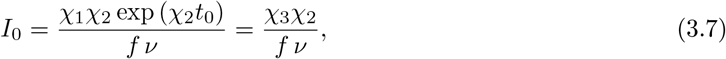

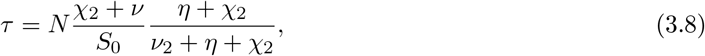

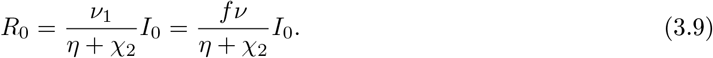

and

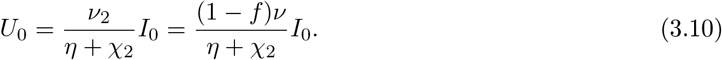

By using the approach described in Diekmann et al [6], van den Driessche and Watmough [25], the basic reproductive number for model (3.1) is given by

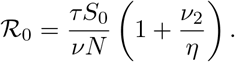

By using (3.8) we obtain

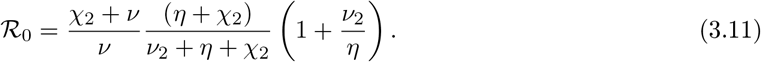

### 3.3 Model SIUR with age structure

In what follows we will denote *N*_1_,…,*N*_10_ the number of individuals respectively for the age classes [0, 10[,…, [90, 100[. The model for the number of susceptible individuals *S*_1_(*t*),…,*S*_10_(*t*), respectively for the age classes [0, 10[,…, [90, 100[, is the following

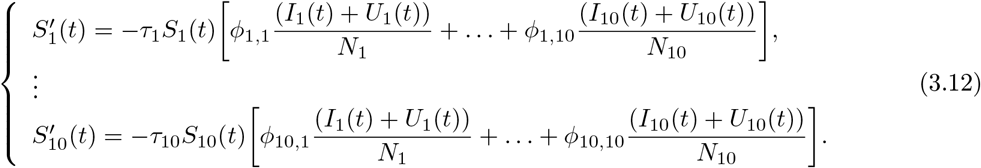

The model for the number of asymptomatic infectious individuals *I*_1_(*t*),…,*I*_10_(*t*), respectively for the age classes [0, 10[,…, [90, 100[, is the following

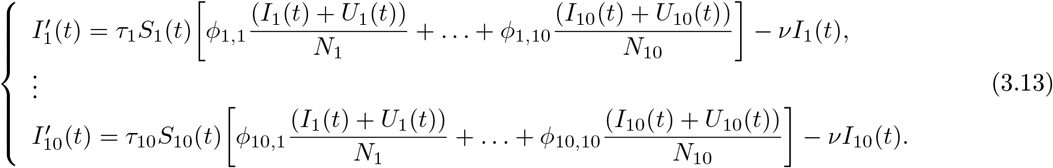

The model for the number of reported symptomatic infectious individuals *R*_1_(*t*),…,*R*_10_(*t*), respectively for the age classes [0, 10[,…, [90, 100[, is

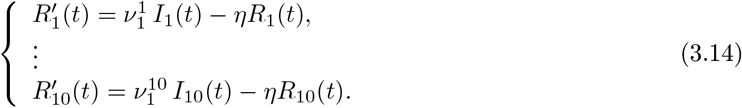

Finally the model for the number of unreported symptomatic infectious individuals *U*_1_(*t*),…,*U*_10_(*t*), respectively in the age classes [0, 10[,…, [90, 100[, is the following

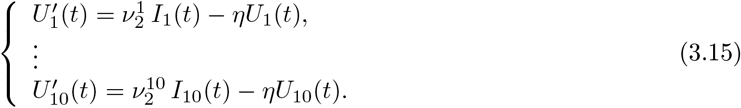

In each age class [0, 10[,…, [90, 100[ we assume that there is a fraction *f*_1_,…,*f*_10_ of asymptomatic infectious individual who become reported symptomatic infectious (i.e. with severe symptoms) and a fraction (1−*f*_1_),…, (1−*f*_10_) who become unreported symptomatic infectious (i.e. with mild symptoms). Therefore we define

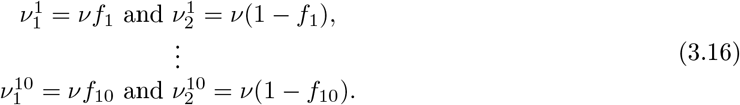

In this model *τ*_1_,…,*τ*_10_ are the respective transmission rates for the age classes [0, 10[,…, [90, 100[.

The matrix *ϕ*_ij_ represents the probability for an individual in the class *i* to meet an individual in the class *j*. In their survey, Prem and co-authors [19] present a way to reconstruct contact matrices from existing data and provide such contact matrices for a number of countries including Japan. Based on the data provided by Prem et al [19] for Japan we construct the contact probability matrix *ϕ*. More precisely, we inferred contact data for the missing age classes [80, 90[ and [90, 100[. The precise method used to construct the contact matrix γ is detailed in Appendix B. An analogous contact matrix for Japan has been proposed by Munasinghe, Asai and Nishiura [18]. The contact matrix γ we used is the following

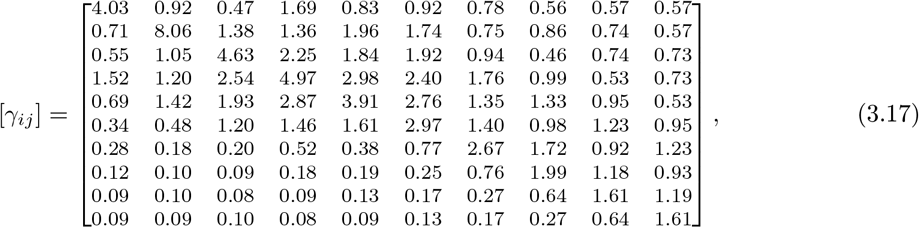

where the *i^th^* line of the matrix *γ_ij_* is the average number of contact made by an individuals in the age class *i* with an individual in the age class *j* during one day. Notice that the higher number of contacts are achieved within the same age class. The matrix of conditional probability *ϕ* of contact between age classes is given by (3.18) and we plot a visual representation of this matrix in Figure 7.

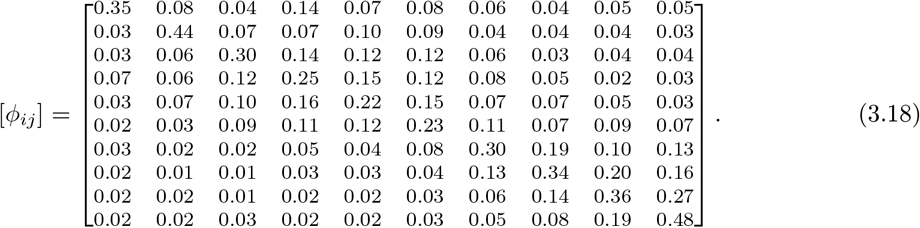

**Figure 7:**
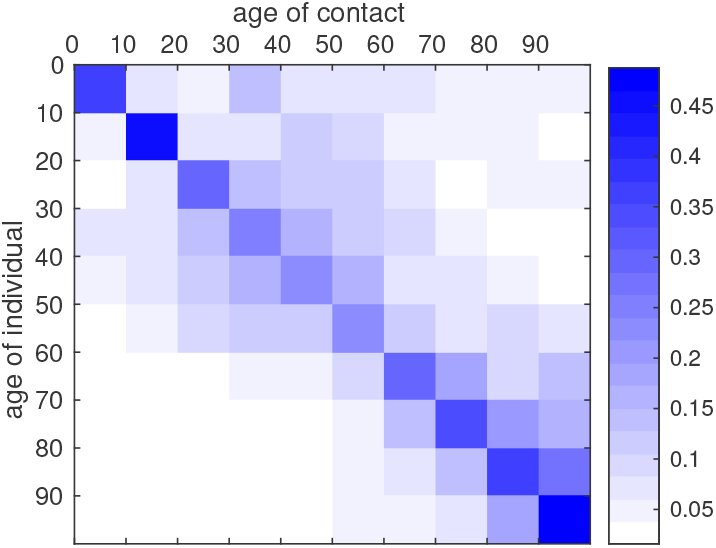
*Graphical representation of the contact matrix ϕ. The intensity of blue in the cell* (*i, j*) *indicates the conditional probability that, given a contact between an individual of age group* i *and another individual, the latter belongs to the age class j. The matrix was reconstructed from the data of Prem et al [19], with the method described in Appendix B*.

## 4 Results

### 4.1 Model without age structure

The daily number of reported cases from the model can be obtained by computing the solution of the following equation:

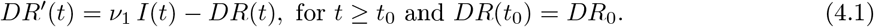

In Figure 8 and Figure 9 we employ the method presented previously in Liu et al [15] to fit the data for Japan without age structure.

**Figure 8:**
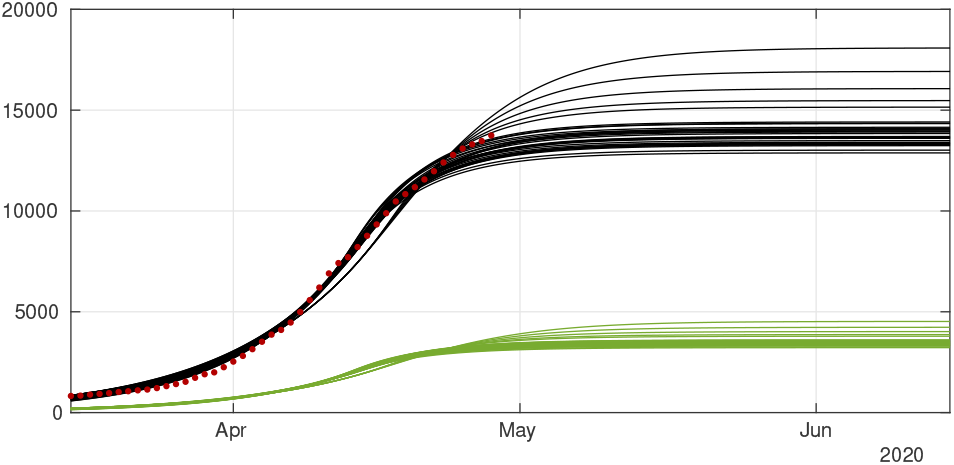
*Cumulative number of cases. We plot the cumulative data (reds dots) and the best fits of the model CR*(*t*) *(black curve) and CU*(*t*) *(green curve). We fix f* =0.8, 1/*η* =7 *days and* 1/*ν* =7 *and we apply the method described in Liu et al [15]. The best fit is d*_1_ = *April* 2*, d*_2_ = *April* 5, D = *April* 27, µ =0.6, χ_1_ = 179, χ_2_ =0.085, χ_3_ =1 *and t*_0_ = *January* 13.

**Figure 9:**
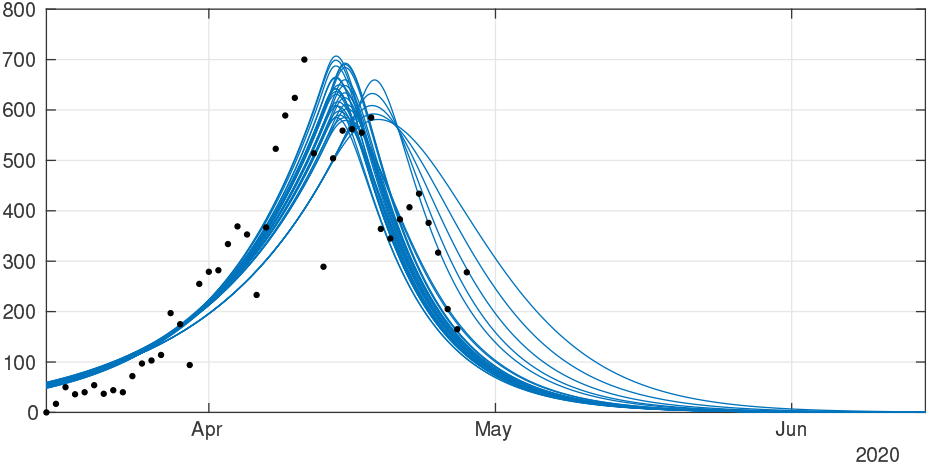
*Daily number of cases. We plot the daily data (black dots) with DR*(*t*) *(blue curve). We fix f* =0.8, 1/η =7 *days and* 1/*ν* =7 *and we apply the method described in Liu et al [15]. The best fit is d*_1_ = *April* 2*, d*_2_ = *April* 5*, N* = *April* 27*, µ* =0.6, χ_1_ = 179, χ_2_ =0.085, χ_3_ =1 *and t*_0_ = *January* 13.

The model to compute the cumulative number of death from the reported individuals is the following

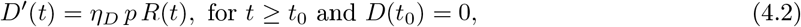

where *η_D_* is the death rate of reported infectious symptomatic individuals and *p* is the case fatality rate (namely the fraction of death per reported infectious individuals).

In the simulation we chose 1/*η_D_* =6 days and the case fatality rate *p* = 0.286 is computed by using the cumulative number of confirmed cases and the cumulative number of deaths (as of April 29) as follows cumulative number of deaths 393

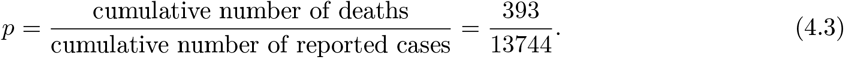

**Figure 10:**
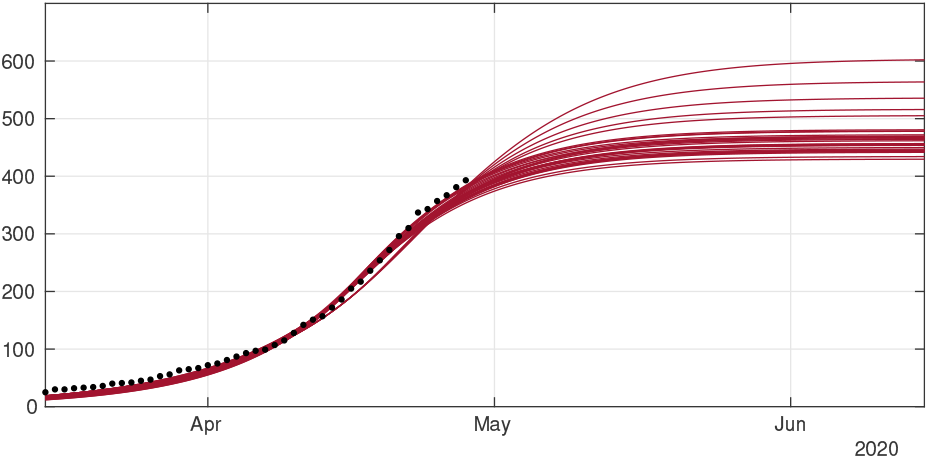
*In this figure we plot the data for the cumulative number of death (black dots), and our best fits for D(t) (red curves)*.

### 4.2 Model with age structure

In order to describe the confinement for the age structured model (3.12)-(3.15) we will use for each age class *i* =1,…, 10 a different transmission rate having the following form

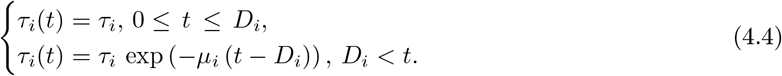

The date *D_i_* is the first day of public intervention for the age class *i* and *µ_i_* is the intensity of the public intervention for each age class.

In Figure 11 we plot the cumulative number of reported cases as given by our model (3.12)-(3.15) (solid lines), compared with reported cases data (black dots). We used the method described in the Appendix A to estimate the parameters *τ_i_* from the data. In Figure 12 we plot the cumulative number of *unreported* cases (solid lines) as given by our model with the same parameter values, compared to the existing data of *reported* cases (black dots).

**Figure 11:**
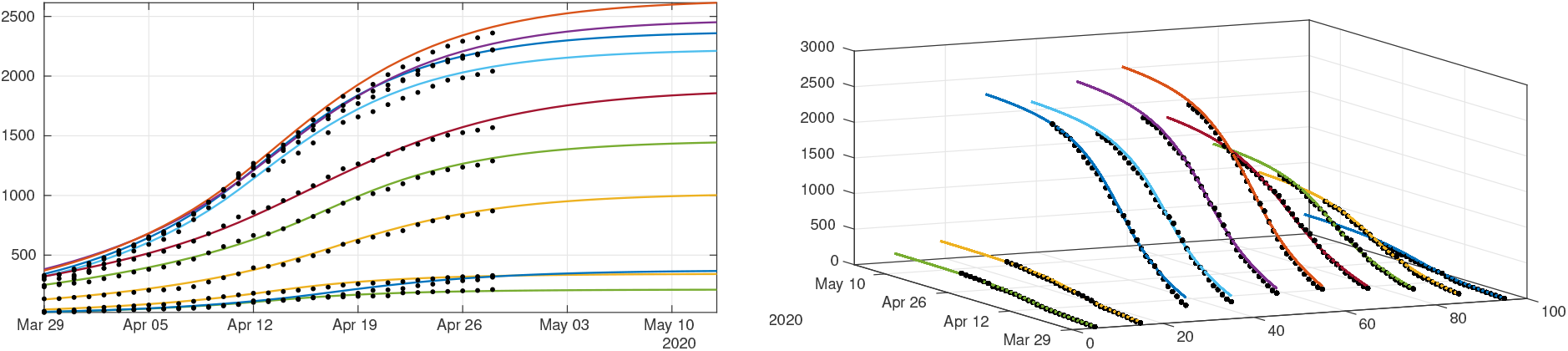
*We plot a comparison between the model (3.12)-(3.15) and the age structured data from Japan by age class. We took* 1/*ν* =1/*η* =7 *days for each age class. Our best fit is obtained for f_i_ which depends linearly on the age class until it reaches 90%, with f*_1_ =0.1*, f*_2_ =0.2*, f*_3_ =0.3*, f*_4_ =0.4*, f*_5_ =0.5*, f*_6_ =0.6*, f*_7_ =0.7*, f*_8_ =0.8*, f*_9_ =0.9*, and f*_10_ =0.9*. The values we used for the first day of public intervention are D_i_* = *Apr. 13 for the 0-20 years age class i* =1, 2*, D_i_* = *Apr. 11 for the age class going from* [20, 30[ *to* [60, 70[ *i* =3, 4, 5, 6, 7*, and D_i_* = *Apr. 16 for the remaining age classes. We fit the data from March* 30 *to April* 20 *to derive the value of 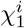 and 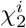 for each age class. For the intensity of confinement we use the values µ*_1_ = *µ*_2_ =0.4829*, µ*_3_ = *µ*_4_ =0.2046*, µ*_5_ = *µ*_6_ =0.1474*, µ*_7_ =0.0744*, µ*_8_ =0.1736*, µ*_9_ = *µ*_10_ =0.1358*. By applying the method described in Appendix A, we obtain τ*_1_ =0.1630*, τ*_2_ =0.1224*, τ*_3_ =0.3028*, τ*_4_ =0.2250*, τ*_5_ =0.1520*, τ*_6_ =0.1754*, τ*_7_ =0.1289*, τ*_8_ =0.1091*, τ*_9_ =0.1211 *and τ*_10_ =0.1642*. The matrix ϕ is the one defined in (3.18)*.

**Figure 12:**
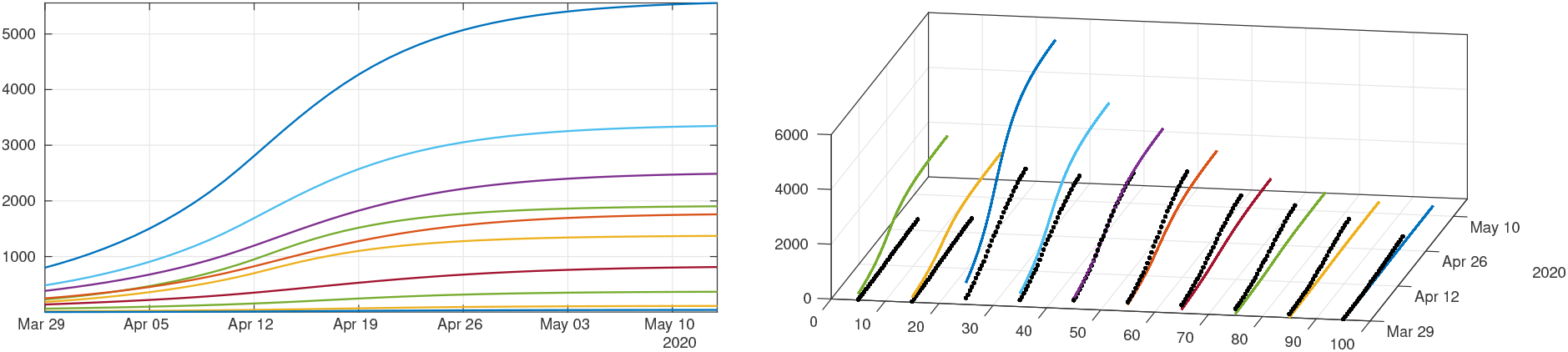
*Cumulative number of unreported cases as given by the fit of the model (3.12)-(3.15) to Japanese data. The solid curves represent the solution of the model and the black dots correspond to the reported cases data. Parameter are the same as in Figure 11*.

**Figure 13:**
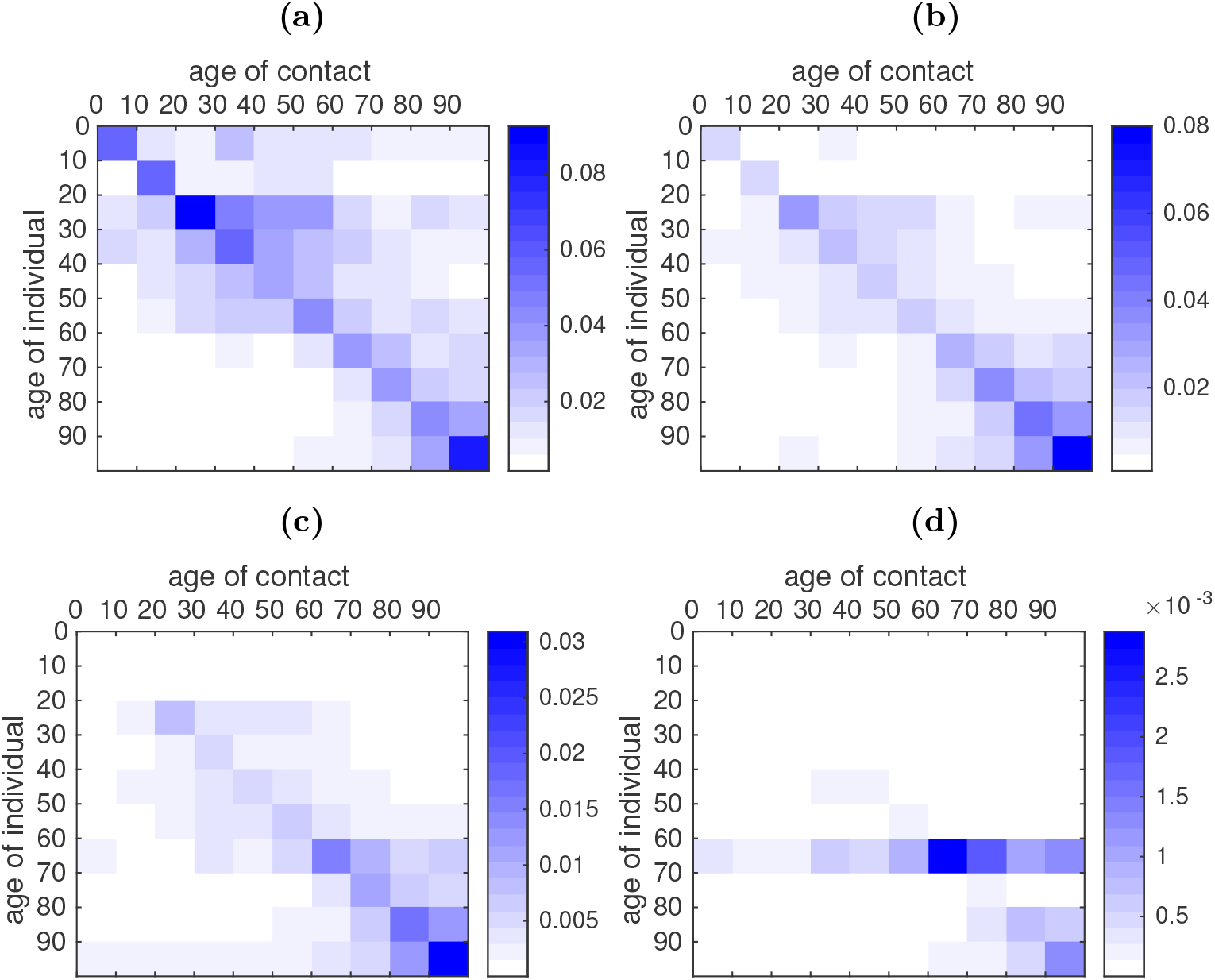
*Rate of contact between age classes according to the fitted data. For each age class in the y-axis we plot the rate of contacts between one individual of this age class and another individual of the age class indicated on the x-axis. (****a****) is the rate of contacts before the start of public measures (April 11). (****b****) is the rate of contacts at the date of effect of the public measures for the last age class (April 16). (****c****) is the rate of contacts one week later (April 23). (****d****) is the rate of contacts one month later (May 16). In this figure we use τ*_1_ =0.1630*, τ*_2_ =0.1224*, τ*_3_ =0.3028*, τ*_4_ =0.2250*, τ*_5_ =0.1520*, τ*_6_ =0.1754*, τ*_7_ =0.1289*, τ*_8_ =0.1091*, τ*_9_ =0.1211 *and τ*_10_ =0.1642*, µ*_1_ = *µ*_2_ =0.4829*, µ*_3_ = *µ*_4_ = 0.2046*, µ*_5_ = *µ*_6_ =0.1474*, µ*_7_ =0.0744*, µ*_8_ =0.1736*, µ*_9_ = *µ*_10_ =0.1358*, and D*1 = *D*2 = *Apr. 13, D*_3_ = *D*_4_ = *D*_5_ = *D*_6_ = *D*_7_ = *Apr. 11, D*_8_ = *D*_9_ = *D*_10_ = *Apr. 16*.

In order to understand the role of transmission network between age groups in this epidemic, we plot in Figure 13 the transmission matrices computed at different times. The transmission matrix is the following

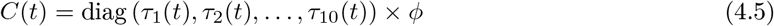

where the matrix *ϕ* describes contacts and is given in (3.18), and the transmission rates are the ones fitted to the data as in Figure 11

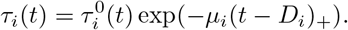

During the early stages of the epidemic, the transmission seems to be evenly distributed among age classes, with a little bias towards younger age classes (Figure 13 (a)). Younger age classes seem to react more quickly to social distancing policies than older classes, therefore their transmission rate drops rapidly (Figure 13 (b) and (c)); one month after the start of social distancing measures, the transmission mostly occurs within elderly classes (60-100 years, Figure 13 (d)).

## 5 Discussion

The recent COVID-19 pandemic has lead many local governments to enforce drastic control measures in an effort to stop its progression. Those control measures were often taken in a state of emergency and without any real visibility concerning the later development of the epidemics, to prevent the collapse of the health systems under the pressure of severe cases. Mathematical models can precisely help see more clearly what could be the future of the pandemic provided that the particularities of the pathogen under consideration are correctly identified. In the case of COVID-19, one of the features of the pathogen which makes it particularly dangerous is the existence of a high contingent of unidentified infectious individuals who spread the disease without notice. This makes non-intensive containment strategies such as quarantine and contact-tracing relatively inefficient but also renders predictions by mathematical models particularly challenging.

Early attempts to reconstruct the epidemics by using SIUR models were performed in Liu et al [11, 12, 13, 14], who used them to fit the behavior of the epidemics in many countries, by including undetected cases into the mathematical model. Here we extend our modeling effort by adding the time series of deaths into the equation. In section 4 we present an additional fit of the number of disease-induced deaths coming from symptomatic (reported) individuals (see Figure 10). In order to fit properly the data, we were forced to reduce the length of stay in the R-compartment to 6 days (on average), meaning that death induced by the disease should occur on average faster than recovery. A shorter period between infection and death (compared to remission) has also been observed, for instance, by Verity et al [26].

The major improvement in this article is to combine our early SIUR model with chronological age. Early results using age structured SIR models were obtained by Kucharski et al [10] but no unreported individuals were considered and no comparison with age-structured data were performed. Indeed in this article we provide a new method to fit the data and the model. The method extends our previous method for the SIUR model without age (see Appendix A).

The data presented in section 2 suggests that the chronological age plays a very important role in the expression of the symptoms. The largest part of the reported patients are between 20 and 60 years old (see Figure 1), while the largest part of the deceased are between 60 and 90 years old (see Figure 5). This suggests that the symptoms associated with COVID-19 infection are more severe in elderly patients, which has been reported in the literature several times (see *e.g*. Lu et al [16], Zhou et al [29]). In particular, the probability of being asymptomatic (our parameter *f*) should in fact depend on the age class.

Indeed, the best match for our model (see Figure 11) was obtained under the assumption that the proportion of symptomatic individual among the infected increases with the age of the patient. This linear dependency of *f* as a function of age is consistent with the observations of Wu et al [28] that the severity of the symptoms increase linearly with age. As a consequence, unreported cases are a majority for young age classes (for age classes less than 50 years) and become a minority for older age classes (more than 50 years), see Figure 12. Moreover, our model reveals the fact that the policies used by the government to reduce contacts between individuals have strongly heterogeneous effects depending on the age classes. Plotting the transmission matrix at different times (see Figure 13) shows that younger age classes react more quickly and more efficiently than older classes. This may be due to the fact that the number of contacts in a typical day is higher among younger individuals. As a consequence, we predict that one month after the effective start of public measures, the new transmissions will almost exclusively occur in elderly classes. The observation that younger ages classes play a major roles in the transmission of the disease has been highlighted several times in the literature, see e.g. Davies et al [5], Cao et al [4], Kucharski et al [10] for the COVID-19 epidemic, but also Mossong et al [17] in a more general context.

We develop a new model for age-structured epidemic and provided a new and efficient method to identify the parameters of this model based on observed data. Our method differs significantly from the existing nonlinear least-squares and statistical inference methods and we believe that it produces high-quality results. Moreover, we only use the initial phase of the epidemic for the identification of the epidemiological parameters, which shows that the model itself is consistent with the observed phenomenon and argues against overfitting. Yet our study could be improved in several direction. We only use reported cases which were confirmed by PCR tests, and therefore the number of tests performed could introduce a bias in the observed data – and therefore our results. We are currently working on an integration of this number of tests in our model. We use a phenomenological model to describe the response of the population in terms of number of contacts to the mitigation measures imposed by the government. This could probably be described more precisely by investigating the mitigation strategies in terms of social network. Nevertheless we believe that our study offers a precise and robust mathematical method which adds to the existing literature.

## Data Availability

The data are accessible from internet and listed in the reference of the paper

## A Appendix: Method to fit of the age structured model to the data

We first choose two days *d*_1_ and *d*_2_ between which each cumulative age group grows like an exponential. By fitting the cumulative age classes [0, 10[,[10, 20[, …and [90, 100[ between *d*_1_ and *d*_2_, for each age class *j* =1,… 10 we can find 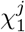 and 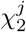

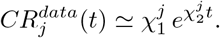

We choose a starting time *t*_0_ ≤ *d*_1_ and we fix

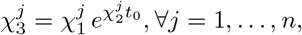

and we obtain

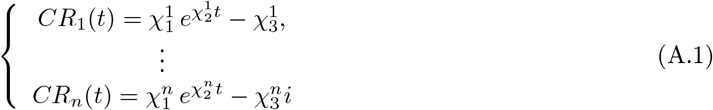

where

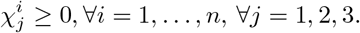

**Figure 14:**
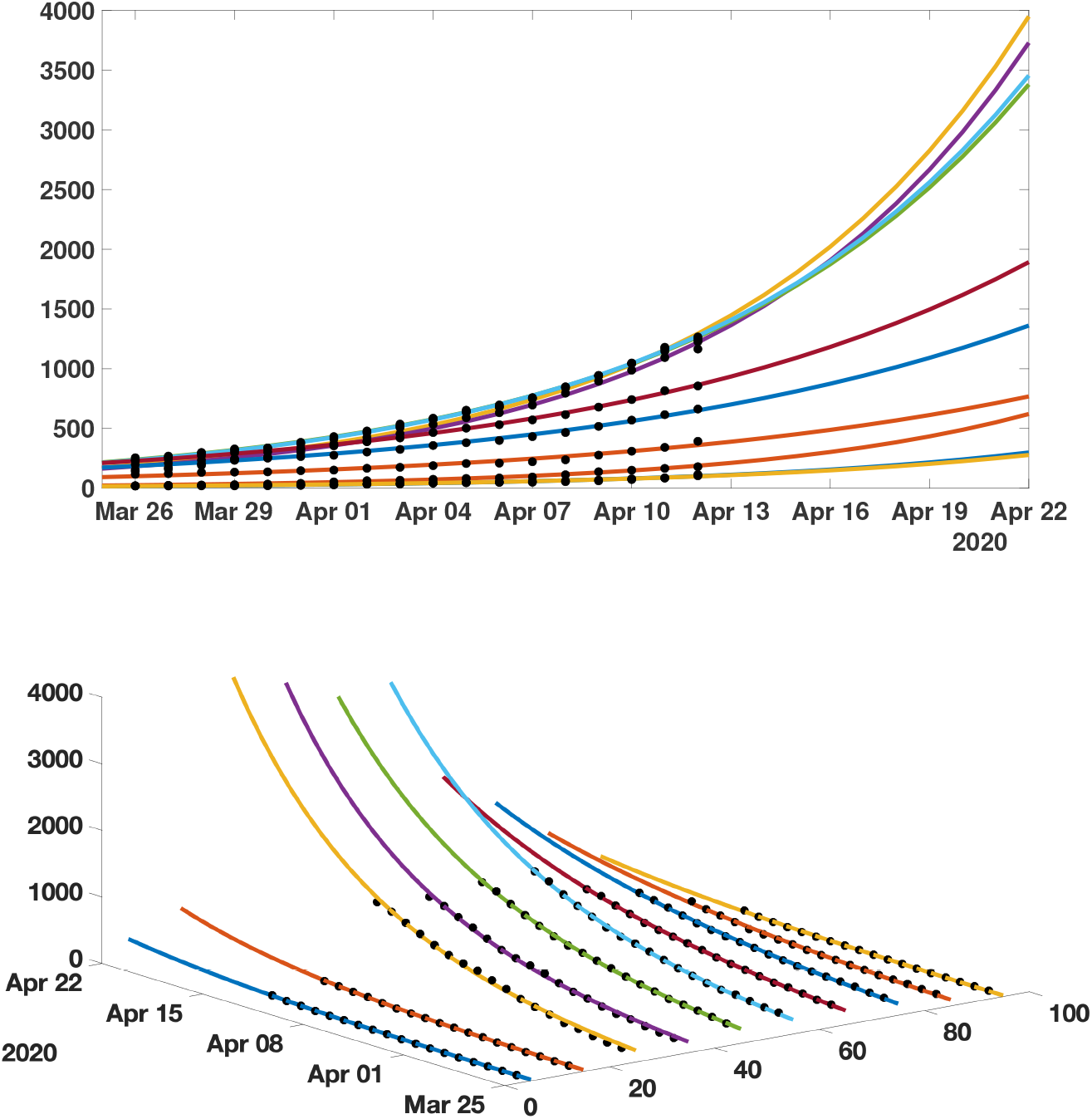
*We plot an exponential fit for each age classes using the data from Japan*.

We assume that

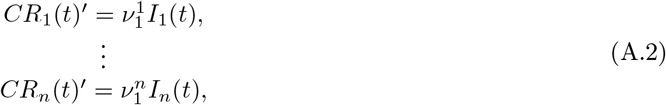

Where

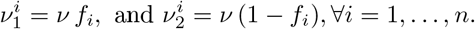

Therefore we obtain

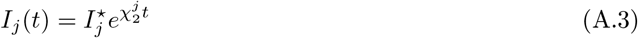

By assuming that the number of susceptible individuals remains constant we have

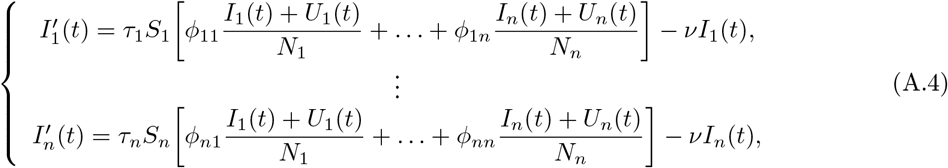

and

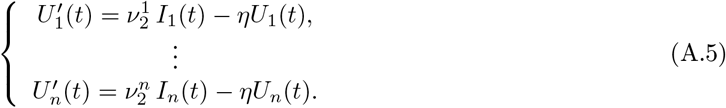

If we assume that the *U_j_*(*t*) have the following form

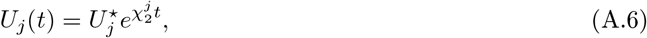

then by substituting in (A.5) we obtain

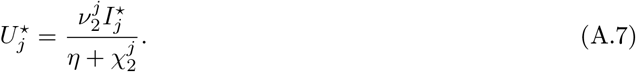

The cumulative number of unreported cases *CU*_j_ (*t*) is computed as

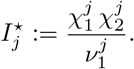

and we used the following initial condition:

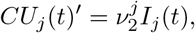

We define the error between the data and the model as follows

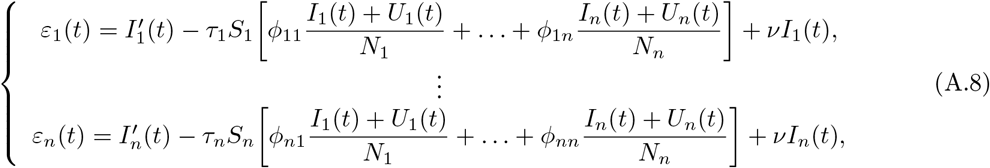

or equivalently

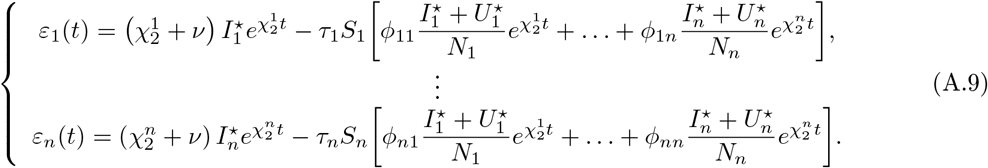

Let the matrix *ϕ* be fixed. We look for the vector τ =(τ_1_,…,τ*_n_*) which minimizes of

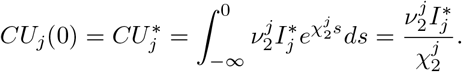

Define for each *j* =1,…,*n*

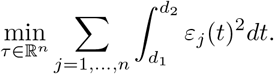

and

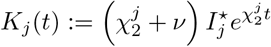

so that

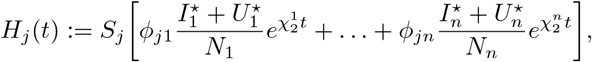

Hence for each *j* =1,…,*n*

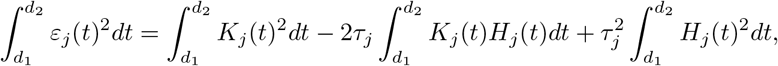

and by setting

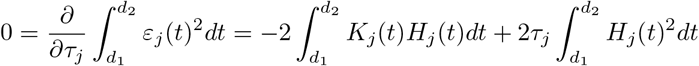

we deduce that

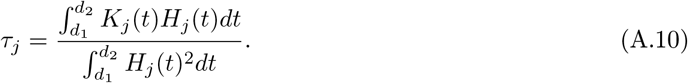

### Remark A.1

*It does not seem possible to estimate the matrix of contact ϕ by using similar optimization method. Indeed, if we look for a matrix ϕ* =(*ϕ_ij_*) *which minimizes*

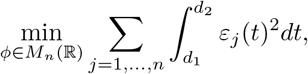

*it turn out that*

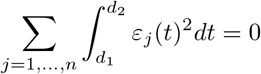

*whenever ϕ is diagonal. Therefore the optimum is reached for any diagonal matrix. Moreover by using similar considerations, if several 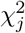 are equal, we can find a multiplicity of optima (possibly with ϕ not diagonal). This means that trying to optimize by using the matrix ϕ does not yield significant and reliable information*.

In the figure 15 below, we present an example of application of our method to fit the Japanese data. We use the period going from March 20 to April 15.

**Figure 15:**
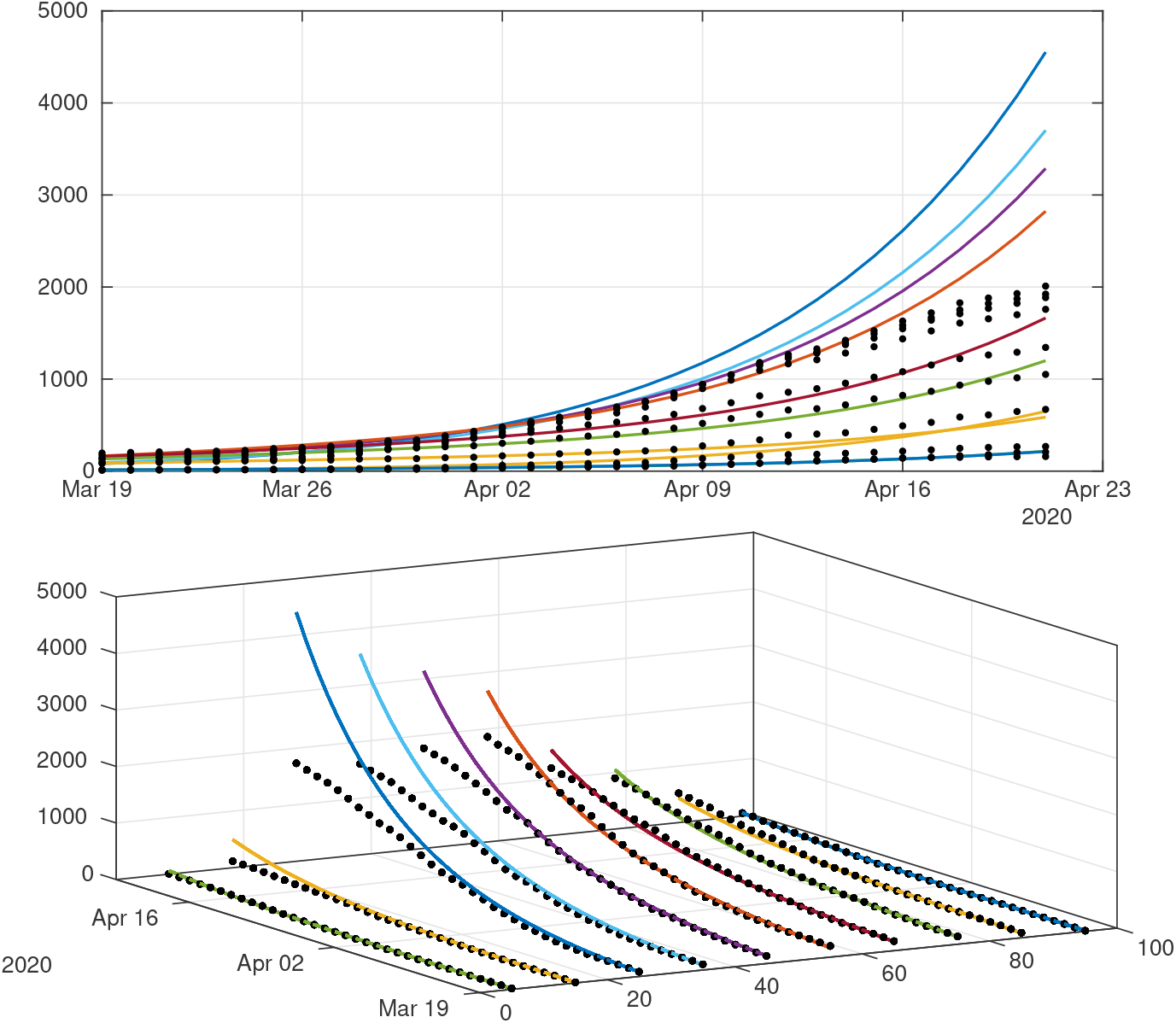
W*e plot a comparison between the model (*3.12*)-(*3.15*) (without public intervention) and the age structured data from Japan. We set* 1/*ν =* 1/*η =* 7 *days, f_i_ which actually depends on the age class, with f*_1_ *=* 0:1*, f*_2_ *=* 0:2*, f*_3_ *=* 0:4*, f*_4_ *=* 0:4*, f*_5_ *=* 0:6*, f*_6_ *=* 0:6*, f*_7_ *=* 0:8*, f*_8_ *=* 0:8*, f*_9_ *=* 0:8*, and f*_10_ *=* 0:9*. and we obtain τ*_1_ *=* 0:1264*, τ*_2_ *=* 0:1655*, τ*_3_ *=* 0:3538*, τ*_4_ *=* 0:2966*, τ*_5_ *=* 0:1513*, τ*_6_ *=* 0:1684*, τ*_7_ *=* 0:1251*, τ*_8_ *=* 0:1168*, τ*_9_ *=* 0:1015*, τ*_10_ *=* 0:1258*. The matrix ϕ is the one defined in (*3.18).

## B Appendix: Construction of the contact matrix

The survey [19] presents reconstructed contact matrices for a number of countries including Japan for the 5-years age classes [0, 5), [5, 10), …, [75, 80) at various locations (work, school, home, and other locations) and a compilation of those contact matrices to account for all locations. The precise description of the compilation is presented in the paper. Note that this paper is a follow-up of Mossong et al. [17] where the survey procedure is described (including the data collection protocol) for several European countries participating in the POLYMOD study.

The data is publicly available online^2^and is presented in the form of a zipped collection of spreadsheets, containing the data for several countries in columns X1 X2 … X16. The columns stand for the average number of contact of one individual of the corresponding age class (0-5 years for X1, 5-10 years for X2, etc…), with an individual of the age class indicated by the row (first row is 0-5 years, second is 5-10 years etc…). Since the age span covered by the study stops at 80, we had to infer the number of contacts for people over the age of 80. We postulated that most people aged 80 or more are retired and that their behaviour does not significantly differs from the behavior of people in the age class [75, 80). Therefore we completed the missing columns by copying the last available information and shifting it to the bottom. We repeated the procedure for lines. We believe that the introduced bias is kept to a minimum since the numerical values are relatively low compared to the diagonal.

Because we use 10-years ages classes and the data is given in 5-years age classes, we had to combine adjacent columns to recover the average number of contacts. To combine columns together, we used the weighted average

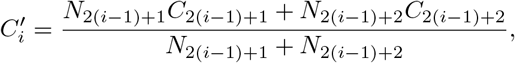

where the column 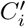 corresponds to the average number of contacts of an individual taken at random in the [10(*i* − 1), 10*i*) and *C_i_* is the average number of contacts of an individual taken at random in the age class [5(*i* − 1), 5*i*). To combine two lines, we simply use the sum of the data

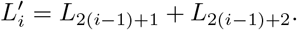

The matrix γ in (3.17) is the transpose of the array obtained by the former procedure applied to the “all locations” dataset. Then *ϕ* is obtained by scaling the lines of γ to 1, *i.e*.

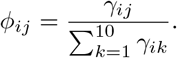

## Acknowledgements

Data from covid19japan.com.

## Footnotes

1 https://covid19japan.com. Accessed May 06, 2020

2 Prem et al [19], Supporting dataset, DOI: 10.1371/journal.pcbi.1005697.s002

